# Cost-Effectiveness of Personalized Policies for Implementing Organ-at-Risk Sparing Adaptive Radiation Therapy in Head and Neck Cancer: A Markov Decision Process Approach

**DOI:** 10.1101/2024.11.05.24316767

**Authors:** Seyedmohammadhossein Hosseinian, Daniel Suarez-Aguirre, Cem Dede, Raul Garcia, Lucas McCullum, Mehdi Hemmati, Aysenur Karagoz, Abdallah S. R. Mohamed, Stephen Y. Lai, Katherine A. Hutcheson, Amy C. Moreno, Kristy K. Brock, Fatemeh Nosrat, Clifton D. Fuller, Andrew J. Schaefer, the Rice/MD Anderson Center for Operations Research in Cancer (CORC), the MD Anderson Head and Neck Cancer Symptom Working Group

## Abstract

**Purpose:** To develop a clinical decision-making model for implementation of personalized organ-at-risk (OAR)-sparing adaptive radiation therapy (ART) that balances the costs and clinical benefits of radiation plan adaptations, without limiting the number of re-plannings per patient, and derive optimal policies for head and neck cancer (HNC) radiation therapy.

**Methods and Materials:** By leveraging retrospective CT-on-Rails imaging data from 52 HNC patients treated at the University of Texas MD Anderson Cancer Center, a Markov decision process (MDP) model was developed to identify the optimal timing for plan adaptations based on the difference in normal tissue complication probability (ΔNTCP) between the planned and delivered dose to OARs. To capture the trade-off between the costs and clinical benefits of plan adaptations, the end-treatment ΔNTCPs were converted to Quality Adjusted Life Years (QALYs) and, subsequently, to equivalent monetary values, by applying a willingness-to-pay per QALY parameter.

**Results:** The optimal policies were derived for 96 combinations of willingness-to-pay per QALY (W) and re-planning cost (RC). The results were validated through a Monte Carlo (MC) simulation analysis for two representative scenarios: (1) W = $200,000 and RC = $1,000; (2) W = $100,000 and RC = $2,000. In Scenario (1), the MDP model’s policy was able to reduce the probability of excessive toxicity, characterized by ΔNTCP ≥ 5%, to zero (down from 0.21 when no re-planning was done) at an average cost of $380 per patient. Under Scenario (2), it reduced the probability of excessive toxicity to 0.02 at an average cost of $520 per patient.

**Conclusions:** The MDP model’s policies can significantly improve the treatment toxicity outcomes compared to the current fixed-time (one-size-fits-all) approaches, at a fraction of their costs per patient. This work lays the groundwork for developing an evidence-based and resource-aware workflow for the widespread implementation of ART under limited resources.

## Introduction

Radiation therapy for head and neck cancer (HNC) has been markedly successful in achieving locoregional tumor control over the past decades, mainly due to the predominance of human papillomavirus associated (HPV+) tumors.^1^ However, a range of acute and chronic toxicities resulting from radiation injuries to non-targeted organs-at-risk (OARs) are still common sequelae in current standard HNC radiation therapy.^2-9^ The risk of normal tissue injury increases with anatomical changes during treatment, such as tumor shrinkage and OAR deformation, which can lead to discrepancies between the planned and actual doses received by the OARs.^10-12^ The principle of adaptive radiation therapy (ART) is to account for these anatomical changes by enabling on-treatment adaptations to the radiation plan (i.e., re-planning).^13-16^

Adapting the radiation plan at each treatment fraction is an ideal form of implementing ART. However, daily re-planning is impractical with current technology because of the finite availability of crucial resources, such as human experts, required for tasks like segmentation and quality assurance. Recent research has aimed to enable a more automated process, for example, by using artificial intelligence (AI)-based algorithms for auto-segmentation.^17-23^ These methods, however, are still developing, and their integration into the clinical workflows is emerging.^24-28^ As a result, the implementation of ART in current practice remains limited to a few treatment fractions. Current institutional guidelines for ART in the US adopt a one-size-fits-all approach and recommend re-planning at fixed intervals, typically once mid-treatment, which fails to consider the uncertain trajectory of toxicities that individual patients may experience.^29,30^ Thus, determining the optimal timing for personalized plan adaptations remains an urgent yet unmet need in HNC radiation therapy to improve care for cancer patients.

This work builds on the previous findings of our research group regarding optimal implementation of OAR-sparing ART under limited resources, as reported by Heukelom et al.^29^ and Nosrat et al.^30^ Heukelom et al.^29^ investigated the optimal timing for ART with a single re-planning allowance, utilizing daily on-treatment CT imaging with a CT-on-rails device. They introduced the difference in normal tissue complication probability (NTCP) between the planned and delivered dose to OARs as an objective selection strategy for ART. Their findings showed that NTCP calculations based on dose differences (ΔNTCP) at fraction 10 were superior to clinical judgment for personalized implementation of ART. Nosrat et al.^30^ investigated the optimal timing for re-planning based on ΔNTCP when multiple adaptations were possible, through a Markov Decision Process (MDP) model. They reported a personalized policy for implementing (OAR-sparing) ART in HNC radiation therapy, with each patient allowed a fixed number of re-plannings.

A critical consideration for successful implementation of ART in practice is its financial feasibility. Nosrat et al.^30^ pioneered applying MDP for personalized ART and successfully identified an optimal re-planning policy for HNC radiation therapy. However, their model assumes the availability of resources for a fixed number of adaptations for every patient. This assumption carries significant financial implications, which may limit the widespread applicability of such policies in clinical settings. Prior research has demonstrated that, while ART can prevent clinically significant toxicities for individual patients, the majority of HNC patients will not require plan adaptations.^29^ Reserving capacity for even a relatively small number of adaptations for each patient could impose a prohibitive financial burden on the healthcare system and create a barrier to the clinical implementation of personalized policies. To address this shortcoming, this paper presents an MDP model for ART decision-making that incorporates financial considerations and seeks an optimal policy that balances the cost and benefits of ART without restricting the number of re-plannings per patient. MDP is a mathematically rigorous framework for decision-making under uncertainty and determining optimal actions in stochastically evolving systems; it has been successfully applied to find optimal timing for various medical interventions.^31-39^ However, its application for decision-making in radiation oncology has largely remained unexplored.^40^

This study serves three specific aims: (1) Develop a clinical decision-making framework for implementing OAR-sparing ART that integrates financial considerations. (2) Determine optimal timing for cost-effective plan adaptations in HNC patients by analyzing data from the University of Texas MD Anderson Cancer Center. (3) Lay the foundation for future research to extend these methodologies to dynamic scenarios, such as online adaptive workflows, with the aim of enabling real-time clinical decision-making.

## Methods and Materials

### Data

This study presents a secondary cost-effectiveness analysis, conducted as per the International Society for Pharmacoeconomics and Outcomes Research (ISPOR) Consolidated Health Economic Evaluation Reporting Standards (CHEERS)^41^. The CHEERS 2022 checklist is provided in **Appendix A (Supplementary Materials)**. We performed *in silico* analysis of a previously reported dataset published by Heukelom et al.^29^ and leveraging a recent Markov Decision Process (MDP) elucidation by Nosrat et al.^30^ which includes data from 52 HNC patients treated with daily CT-on-Rails Image-Guided Radiation Therapy (IGRT) at the University of Texas MD Anderson Cancer Center between 2007 and 2013. Patients were treated with daily kilovoltage CT imaging, and daily CT-image-based replanning was generated to assess “virtual daily replanning” across all-delivered fractions, assuming a “fixed GTV/CTV1” approach (i.e., normal-tissue and weight loss were accounted for, but GTV and high-dose CTV volumes were neither altered nor dose-reduced). This secondary analysis was conducted under institutional review board approval MDA RCR03-0800. These patients received radiation therapy (9-beam step-and-shoot IMRT), either alone or in combination with chemotherapy or cetuximab. The primary cancer sites included oropharynx, nasopharynx, sinonasal region, oral cavity, and larynx. In this cohort, 69% of the patients were male, and 31% were female. The patient characteristics are provided in **Appendix B (Supplementary Materials)**.

To develop a novel MDP model for cost-effective ART implementation, we utilized the published analysis results of Heukelom et al.^29^ and Nosrat et al.^30^ on this dataset. Specifically, we leveraged the ΔNTCP calculations of Heukelom et al.^29^ along with the probabilistic estimates of ΔNTCP trajectories from Nosrat et al.^30^ For each patient in this cohort, Heukelom et al.^29^ analyzed the deviation of the delivered (accumulated) dose from the planned dose on a daily basis for multiple OARs and, accordingly, calculated ΔNTCP for xerostomia, dysphagia, parotid gland dysfunction, and tube feeding dependency at 6 months post-treatment. Building on these results, Nosrat et al.^30^ developed an MDP model, capturing the patient’s state of toxicity by ΔNTCP, and calculated the associated transition probabilities. In a similar manner, we consider ΔNTCP as our decision-making criterion and use the transition probabilities reported by Nosrat et al.^30^ to develop an MDP model that balances clinical benefits with the cost of plan adaptations. The ΔNTCP values and associated transition probabilities from this dataset are presented in **Appendix C (Supplementary Materials)**.

### Decision Model

The MDP model captures the state of toxicity through ΔNTCP. Before treatment begins, the system is at ΔNTCP = 0%, indicating that the expected post-treatment toxicities align with the treatment plan. Throughout the treatment, the patient’s ΔNTCP follows a stochastic trajectory due to uncertain anatomical changes. This uncertainty is captured by transition probabilities, which quantify the likelihood of changes in ΔNTCP from one epoch (e.g., treatment fraction) to the next. At each epoch, the clinician observes the system’s state (i.e., ΔNTCP) and may choose between two actions: (1) re-plan, or (2) continue with the current plan. Re-planning, which incurs a monetary cost, modifies the probabilistic transition towards more favorable outcomes/states. At the end of the treatment, the process culminates in a terminal reward, determined by the system’s final state (i.e., the patient’s end-treatment ΔNTCP).

To capture the trade-off between the cost and clinical benefits of plan adaptations, the terminal reward in our model is defined as the *monetary* equivalent of the clinical benefits. This is achieved by converting end-treatment ΔNTCPs to Quality Adjusted Life Years (QALYs) and then applying a *willingness-to-pay per QALY* parameter to determine the monetary equivalent of the QALY gains. An optimal solution to this MDP determines the best action (to re-plan or not) at each decision epoch based on the patient’s toxicity state (ΔNTCP). Such an optimal policy (i.e., the set of optimal actions) will depend on the re-planning cost and clinical benefits.

Components of the MDP model, developed using the dataset described previously, are as follows:

### Decision epochs

Given a typical treatment period of 33-35 fractions for HNC, decision epochs were set at fractions 10, 15, 20, and 25 (weekly). Fraction 5 was excluded based on earlier research showing that anatomical changes are unlikely to occur early during treatment.^29^ Similarly, fraction 30 was omitted due to its proximity to the treatment’s conclusion, where adjustments would have minimal effect on the overall dose delivered to OARs. This choice of decision epochs follows the MDP model of Nosrat et al.^30^

### States

At each decision epoch, the state of the system was represented by ΔNTCP, ranging from 0% to 12%, based on the findings of Heukelom et al.^29^ The results of Heukelom et al.^29^ and the transition probability calculations of Nosrat et al.^30^ are based on the number of patients exhibiting a certain ΔNTCP for *any* of the considered toxicities (i.e., xerostomia, dysphagia, parotid gland dysfunction, and tube feeding dependency). Consequently, the definition of the states in our model adopts a holistic approach, aiming to protect against all these toxicities.

### Actions

At each state, two possible actions were included: (1) re-planning, or (2) continuing with the current plan (no re-planning).

### Transition probabilities

The stochastic transition of the system’s state (i.e., ΔNTCP) from one decision epoch to the next is governed by transition probabilities, as a function of the action taken. We leveraged the transition probabilities calculated by Nosrat et al.^30^ in our model.

### Immediate rewards

In the MDP setting, each action at each state may yield an immediate reward. We note that “reward” is standard terminology in the MDP literature. In our application, the immediate reward is the cost of re-planning (only associated with the action “re-planning”), which is represented as a negative number. The cost of re-planning may vary across institutions. To ensure a comprehensive analysis, we considered various costs in our model, ranging from $500 to $2000 (in $100 increments) per re-planning, based on recently reported re-planning costs in the literature.^42,43^

### Terminal rewards

The terminal reward of the process was calculated based on the terminal state (i.e., end-treatment ΔNTCP), as follows:

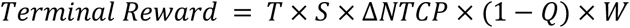

where *T* denotes the number of years (post-treatment) that the patient’s quality of life is considered, *S* is the patient’s *T*-year survival probability, *Q* represents a quality-of-life factor used to transform end-treatment ΔNTCP into changes in QALY, which is determined by the type and severity of toxicities, and *W* denotes the willingness-to-pay per QALY (in $). Details of the derivation of this formula are presented in **Appendix D (Supplementary Materials)**. In our analysis, we used *T=* 5 years and *S =* 0.685, as the 5-year overall survival rate of HNC patients.^44^ The literature reports a range of values for *Q* concerning different radiation toxicities in HNC patients;^45^ see **Appendix E (Supplementary Materials)**. Following our holistic approach for protecting against the toxicities considered by Heukelom et al.^29^, we used *Q =* 0.80 in our analysis. The literature reports various values for willingness-to-pay per QALY as well.^40,46^ To ensure a comprehensive analysis, we used a range of values for *W*, including $50,000, $75,000, $100,000, $150,000, $200,000, and $250,000.

Like immediate rewards, terminal rewards in our application have negative values because ΔNTCP represents the extent to which the actual NTCP is worse than the planned NTCP. The objective of MDP is to find a policy that maximizes the sum of immediate and terminal rewards. Since the rewards are negative in our application, this can be viewed as minimizing the patient’s total loss, that is the combination of the monetary equivalent of the patient’s loss in the quality of life and the money spent on re-planning.

The MDP model was implemented in Python and solved using the *mdptoolbox* library.^47^ The code and related results are publicly available in the online repository of this project.

## Results

An optimal policy was calculated for each pair of willingness-to-pay per QALY (W) and re-planning cost (RC). Therefore, a total of 6 × 16 = 96 optimal policies were obtained. For each W-RC pair, the optimal policy indicates the optimal action (0 for “no re-planning” and 1 for “re-planning”) for each ΔNTCP state at each decision epoch. **Table 1** demonstrates the optimal policy for W = $200,000 and RC = $1,000, as an example. Since the presentation of the optimal policies (for all 96 cases) is extensive and clinically less informative, we present them based on the concept of *re-planning ΔNTCP threshold*, as follows. The optimal policies in their extensive form are available in the project’s online repository.

**Table 1:** Optimal policy for W = $200,000 and RC = $1,000. An entry 1 in the optimal policy indicates that the optimal action (at the corresponding treatment fraction and ΔNTCP) is “re-planning” and an entry 0 indicates the optimal action is “no re-planning.”

| | $\Delta NTCP$ | | | | | | | | | | | | |
| --- | --- | --- | --- | --- | --- | --- | --- | --- | --- | --- | --- | --- | --- |
| Decision Epoch | 0% | 1% | 2% | 3% | 4% | 5% | 6% | 7% | 8% | 9% | 10% | 11% | 12% |
| Fraction 10 | 0 | 0 | 0 | 1 | 1 | 1 | 1 | 1 | 1 | 1 | 1 | 1 | 1 |
| Fraction 15 | 0 | 0 | 0 | 0 | 1 | 1 | 1 | 1 | 1 | 1 | 1 | 1 | 1 |
| Fraction 20 | 0 | 0 | 0 | 1 | 1 | 1 | 1 | 0 | 1 | 1 | 1 | 1 | 1 |
| Fraction 25 | 0 | 0 | 0 | 0 | 1 | 1 | 0 | 1 | 1 | 1 | 1 | 1 | 1 |

To obtain clinically interpretable policies, for each optimal policy obtained from the MDP model, we considered the smallest ΔNTCP value that justified re-planning as the *re-planning threshold*. This converts an optimal policy to a *threshold policy* that recommends re-planning only if ΔNTCP is greater than or equal to the threshold. For example, ΔNTCP = (3%, 4%, 3%, 4%$) at the treatment fractions 10, 15, 20, and 25, respectively, are the decision-making thresholds associated with the optimal policy presented in **Table 1**.

Figure 1. summarizes the threshold policies for all the W-RC pairs considered. Each subfigure corresponds to a willingness-to-pay per QALY value, with each row representing a re-planning cost. For each W-RC pair, the policy is depicted by four ΔNTCP thresholds for decision-making at treatment fractions 10, 15, 20, and 25 (i.e., decision epochs). The threshold policy associated with the optimal policy of **Table 1** is highlighted with a red box in this figure.

**Figure 1.**
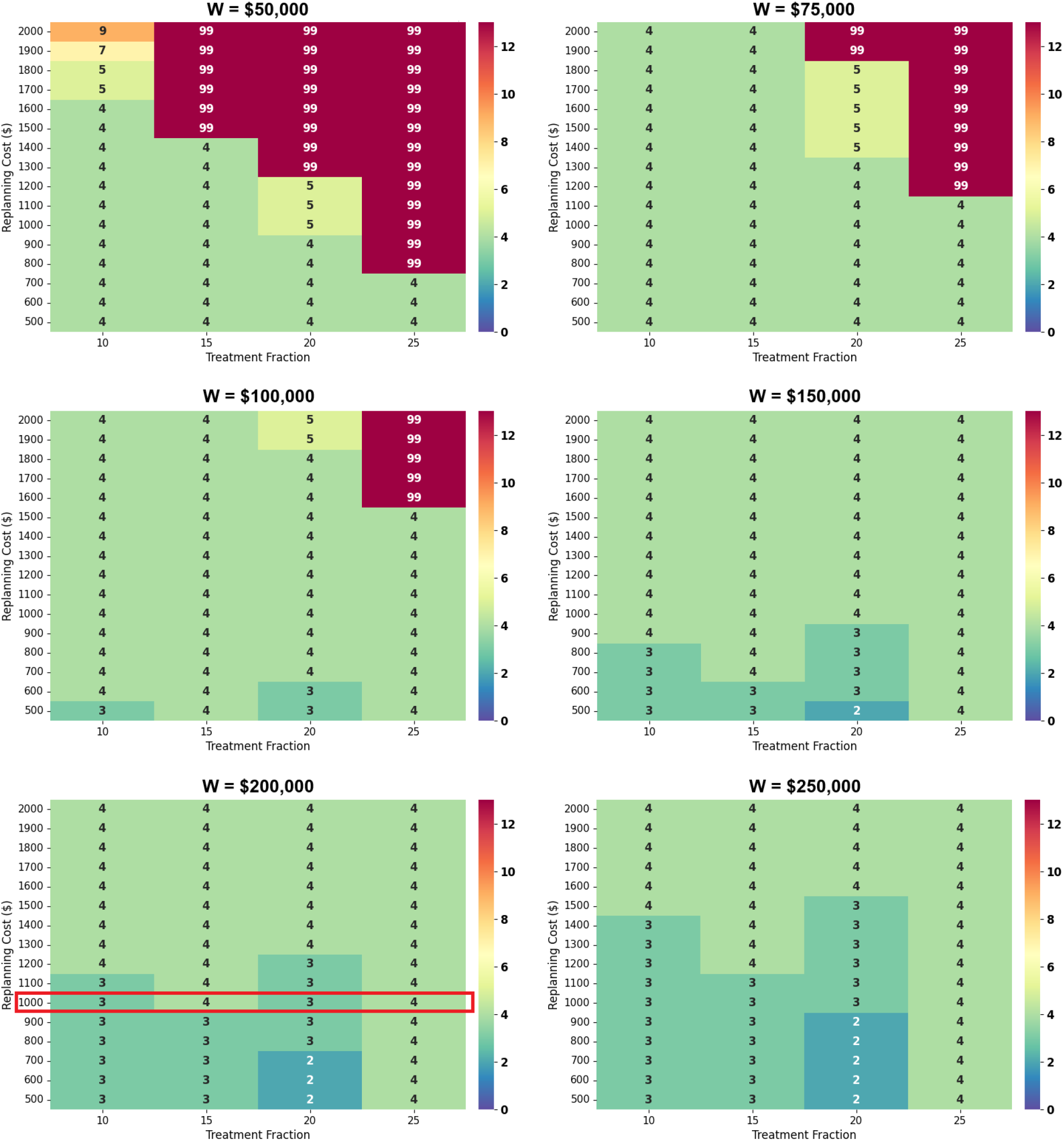
ΔNTCP (%) thresholds for re-planning at each decision epoch. Each subfigure concerns a fixed value of willingness-to-pay per QALY (W) and presents ΔNTCP thresholds for various re-planning costs (RC). The threshold policy associated with the optimal policy of *Table 1* is highlighted with a red box. The threshold of ΔNTCP = 99% in this figure implies “no re-planning” for any ΔNTCP value.

### Model Validation

To validate these results, we conducted a Monte Carlo (MC) simulation study to compare the clinical and financial outcomes of the MDP model’s threshold policies against two fixed-time (one-size-fits-all) policies. MC simulation is an established method for cost-effectiveness analysis of medical interventions; it provides probabilistic estimates of the cost and utility of a specific policy and enables the comparison of different policies in these terms.^40^ In our analysis, we considered three re-planning policies: (a) the patient receives a single “re-planning” at fraction 10, regardless of their toxicity status; (b) the patient receives a single “re-planning” at fraction 15, regardless of their toxicity status; (c) the threshold policy of the MDP model, which does not limit the number of re-plannings, is followed. Since the threshold policies depend on W and RC, we examined two representative scenarios with respect to these parameters: (1) W = $200,000 and RC = $1,000; (2) W = $100,000 and RC = $2,000. These scenarios are intended to represent different conditions regarding economic prosperity and resource availability. The MC analysis involved 10,000 simulation runs for each policy.

Figure 2. illustrates the distribution of end-treatment ΔNTCP for policies (a), (b), and (c) with W = $200,000 and RC = $1,000, as well as the case where no re-planning is done for the patient, referred to as policy (d). When policy (d) was followed, meaning no re-planning at all, the expected end-treatment ΔNTCP (mean) was 2.36% (standard deviation (SD) = 2.97%). Following policies (b) and (a)—a single re-planning at fractions 15 and 10, respectively—the expected end-treatment ΔNTCP reduced to 1.57% (SD = 1.92%) and 1.20% (SD = 1.48%), respectively. The MDP model’s threshold policy, i.e., policy (c), led to the expected end-treatment ΔNTCP of 0.98% (SD = 0.82%).

**Figure 2.**
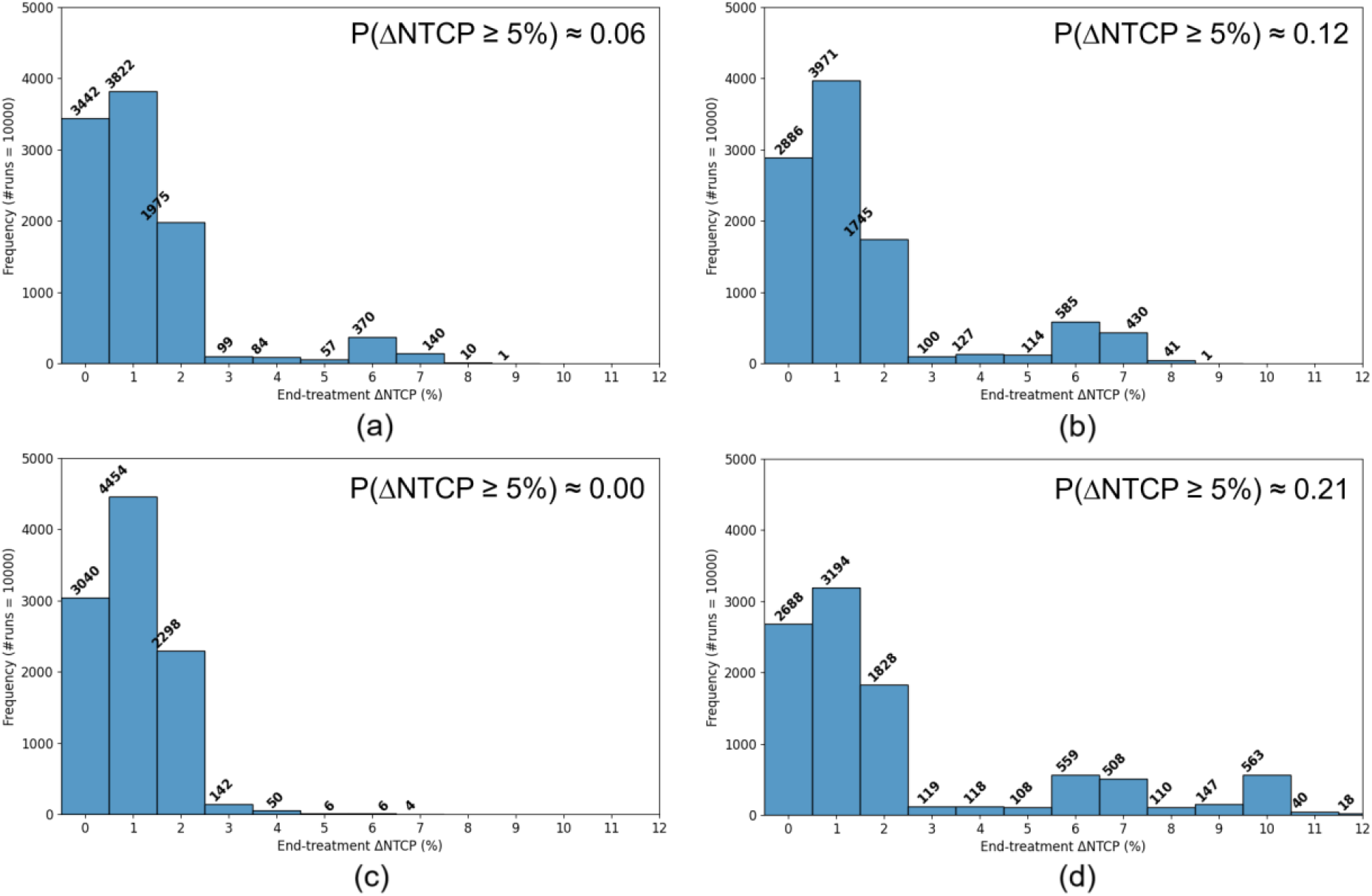
Distribution of the end-treatment ΔNTCP (%) under the following policies: (a) the patient receives a single “re-planning” at fraction 10; (b) the patient receives a single “re-planning” at fraction 15; (c) the threshold policy of the MDP model for W = $200,000 and RC = $1,000 is followed; (d) no re-planning at all. The number of simulation runs was 10,000.

As highlighted by Heukelom et al.^29^ the majority of HNC patients in this cohort did not require re-planning, which is manifested through the small mean ΔNTCPs, even under policy (d). However, some patients in this cohort exhibited considerable discrepancies between the planned and actual dose to OARs, resulting in large ΔNTCP values (up to 12%). In this regard, we examined the effectiveness of the threshold policy in preventing large ΔNTCPs. Under policy (d), i.e., no re-planning, the probability of the patient experiencing ΔNTCP ≥ 5% was 0.21. This probability reduced to 0.12 and 0.06 when the policy was to re-plan at fractions 15 and 10, i.e., policies (b) and (a), respectively. Under the MDP model’s threshold policy, i.e., policy (c), the probability of ΔNTCP ≥ 5% was approximately zero.

Regarding the financial aspect, policies (a) and (b) require re-planning once, regardless of the patient’s toxicity status. Therefore, the number of re-planning under these policies is one, implying an average cost of $1,000 per patient for this scenario. The outcome of the MC analysis under the MDP model’s threshold policy is a distribution over all possible number of re-plannings, ranging from 0 to 4 (re-planning at all four decision epochs), which is illustrated in **Figure 3**. The expected number of re-plannings (mean) under this policy was 0.38 (SD = 0.68), implying an average cost of $380 per patient.

**Figure 3.**
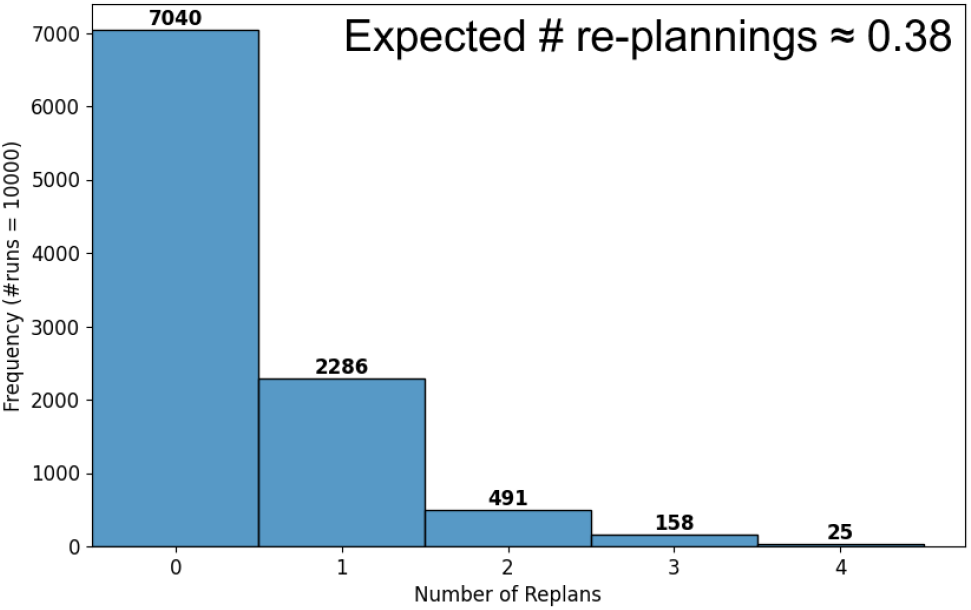
Distribution of the number of re-plannings under the threshold policy of the MDP model for W = $200,000 and RC = $1,000. The number of simulation runs was 10,000.

Since the MDP model’s threshold policies depend on the willingness-to-pay per QALY and re-planning cost, we repeated the analysis with a different set of values for these parameters; that is W = $100,000 and RC = $2,000. This second scenario resembles lower-income communities with greater resource constraints compared to the previous scenario. **Figure 4** illustrates the distribution of end-treatment ΔNTCP along with the distribution of the number of re-plannings when the threshold policy of the MDP model with these parameters was followed. The expected end-treatment ΔNTCP under the threshold policy for this scenario was 1.14% (SD = 1.12%). The probability of experiencing ΔNTCP ≥ 5% was 0.02. The expected number of re-plannings for this scenario was 0.26 (SD = 0.56), implying the average cost of $520 per patient.

**Figure 4.**
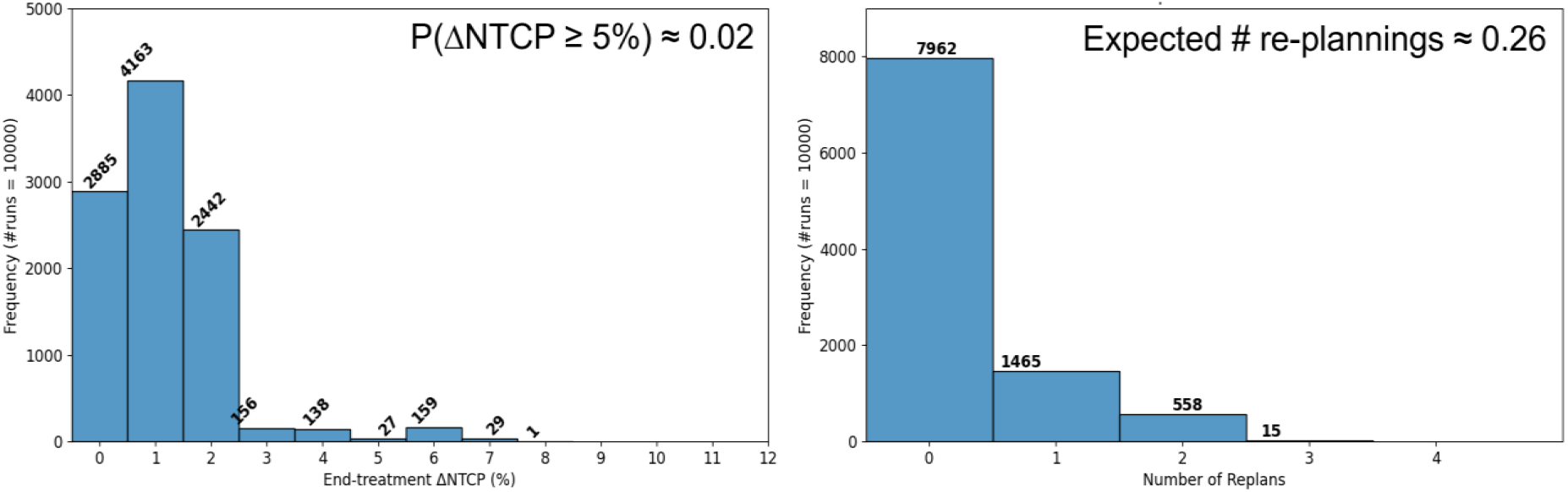
Distribution of the end-treatment ΔNTCP (left) and distribution of the number of re-plannings (right) under the threshold policy of the MDP model for W = $100,000 and RC = $2,000. The number of simulation runs was 10,000.

The MC simulation results are summarized in **Table 2**.

**Table 2:** Summary of the Monte Carlo simulation results. The willingness-to-pay per QALY and re-planning cost are denoted by W and RC, respectively.

| Policy | Clinical Outcome |  | Financial Outcome |  |
| --- | --- | --- | --- | --- |
| | Expected end-treatment ΔNTCP | Probability ΔNTCP ≥ 5% | Expected number of re-plannings | Average cost per patient |
| **No re-planning** | 2.36% (SD = 2.97%) | 0.21 | 0.00 | \$0 |
| **A single re-planning at fraction 15** | 1.57% (SD = 1.92%) | 0.12 | 1.00 | Scen.1: \$1,000<br>Scen.2: \$2,000 |
| **A single re-planning at fraction 10** | 1.20% (SD = 1.48%) | 0.06 | 1.00 | Scen.1: \$1,000<br>Scen.2: \$2,000 |
| **MDP threshold policy: Scenario 1**<br>**$W = \$200,000$ and $RC = \$1,000$** | 0.98% (SD = 0.82%) | 0.00 | 0.38 | \$380 |
| **MDP threshold policy; Scenario 2**<br>**$W = \$100,000$ and $RC = \$2,000$** | 1.14% (SD = 1.12%) | 0.02 | 0.26 | \$520 |

## Discussion

The MC simulation analysis results demonstrate that the MDP model’s threshold policies outperform the fixed-time policies in both clinical and financial outcomes. As noted by Heukelom et al.^29^ most patients in this cohort did not require re-planning due to minimal anatomical changes. Consequently, the expected end-treatment ΔNTCP for all the considered policies falls within a narrow range, potentially masking the significance of the benefits of the MDP model’s threshold policies compared to the fixed-time policies. The main advantage of the MDP model’s threshold policies lies in their ability to prevent excessive toxicities. A subset of patients in this cohort did experience notable anatomical changes and large ΔNTCPs. The analysis indicates that a single re-planning at fraction 15 could approximately halve the probability of patients experiencing ΔNTCP ≥ 5% (from 0.21 to 0.12). If re-planning occurs earlier at fraction 10, this probability drops to less than one third (from 0.21 to 0.06). Following the MDP model’s threshold policy for Scenario 2 reduces the probability to under one tenth (from 0.21 to 0.02), while the threshold policy for Scenario 1 makes it negligible (from 0.21 to 0.00). The choice of ΔNTCP ≥ 5% in our analysis is motivated by the threshold of ΔNTCP = 5% set by the Dutch Society for Radiation Oncology for assigning patients to proton therapy for grade III complications.^29^ Finally, we note that these results are aligned with the findings of Heukelom et al.^29^ regarding the clinical advantages of re-planning at fraction 10 compared to fraction 15.

The average cost per patient under each policy demonstrates the financial advantage of the MDP model’s threshold policies. The MDP model’s policies allocate re-planning resources to patients who need them most, without limiting the number of re-plannings per patient, in contrast to the fixed-time policies that imply re-planning for every patient. Since many patients are expected to experience minimal anatomical changes, the expected number of re-plannings per patient under the MDP model’s policy for Scenario 1 was 0.38. Given the re-planning cost of $1,000 in this scenario, this translates to the average cost of $380 per patient—almost one third of the cost incurred if every patient received one re-planning. For Scenario 2, the expected number of re-plannings per patient was 0.26 under the MDP model’s threshold policy, which translates to the expected cost of $520 per patient—nearly a quarter of the cost of fixed-time policies. This result is of a significant importance for financial stakeholders (e.g., insurers) because harnessing the full capacity of ART to improve patient care requires flexible re-planning as needed—not limited to one or two instances—and the MDP analysis demonstrates that this can be achieved with an average of less than one re-planning per patient. Finally, we note that willingness-to-pay per QALY is a measure of the population’s economic prosperity rather than an individual patient’s willingness to pay for their treatment.

As noted earlier, the optimal policies of the MDP model were not necessarily threshold policies. This complicates their clinical interpretation and makes them less amenable to implementation in practice. To address this, we converted the optimal policies into threshold policies by defining the threshold as the smallest ΔNTCP that justifies re-planning. It is important to recognize that the MDP model’s optimal policies are highly dependent on transition probabilities, which are often inferred from limited-size datasets and can be susceptible to noise. This can result in counterintuitive recommendations, such as “no re-planning” at fraction 20 for ΔNTCP = 7% in the optimal policy shown in **Table 1**, while the recommendation for ΔNTCP between 3% and 6% as well as ΔNTCP greater than or equal to 8% at this fraction are “re-planning.” This discrepancy stems from the transition probabilities calculated based on a cohort of only 52 patients. Thus, the threshold policies not only enhance the clinical interpretability and applicability of the policies derived from the MDP analysis but also incorporate clinical intuition to mitigate the noise inherent in small datasets.

The presented findings are subject to the limitations inherent to a single-institution retrospective *in silico* analysis. The results are constrained by the inferred transition probabilities based on data gathered from 52 patients. While our dataset is among the most curated and well-accredited datasets relevant to adaptive radiation therapy for HNC, we recognize the significance of sample size for MDP calibration and acknowledge the need for external validation with larger datasets. These findings are also constrained by the limitations of the ΔNTCP calculations and the inference of the transition probabilities, elaborated in Heukelom et al.^29^ and Nosrat et al.^30^ Particularly, the dataset did not include adaptations or daily re-optimization *in vivo*, and the daily dose accumulation was calculated post hoc from high-resolution imaging data. As a result, there were discrepancies, which were omitted, leading to gaps in the resulting NTCPs. As the dataset did not include patients’ records for the entire treatment period, transition probabilities for the missing time points were inferred from available data in conjunction with clinical judgment. The results are also specific to the toxicities considered and the NTCP models used. Novel approaches to NTCP modeling, such as cluster-based methods^8^, can potentially lead to improved policies. Finally, the results are subject to potential inaccuracies and limitations regarding the values obtained from the literature, particularly the parameters used to calculate the monetary equivalent of the clinical gains. Thus, these findings should be considered an informative semi-synthetic use case rather than a definitive basis for large-scale implementation of ART.

The results presented in this paper utilized CT-on-rails volumetric IGRT data for HNC. However, the proposed methodology is applicable to various platforms, notably MR-linacs that offer a compelling use case, as well as other cancer sites. The decision epochs in our model correspond to weekly re-planning intervals, reflecting our current institutional adaptive protocols,^1^ but the methodology can be scaled to daily treatment decisions. The model is flexible for use in both resource-rich and resource-limited facilities and is capable of considering the economic and financial standing of the communities they serve. Additionally, the model allows for further treatment personalization, such as incorporating stratified survival probabilities (e.g., HPV+ vs. HPV-tumors) and considering different timeframes for accounting for patients’ quality of life. The presented model focuses on OAR-sparing ART, assuming that the prescribed dose to the tumor remains unchanged during treatment. Incorporating dose adaptations for the tumor and considering changes in tumor control probability (TCP) during treatment presents a promising avenue for future research. Our results employ ΔNTCP as an evidence-based measure of treatment toxicity. We recognize that this setting adds a layer of computational complexity to clinical decision-making. This motivates future research to leverage the presented framework for decision-making based on easily observable treatment outcomes through partially observable Markov decision processes (POMDPs).^48^

## Conclusion

This paper presents the first MDP model for implementing *patient-specific* OAR-sparing ART in HNC that balances the costs and benefits of plan adaptations and renders cost-effective personalized policies. By analyzing data collected from 52 patients treated at MD Anderson Cancer Center with high-quality CT-based replanning, we derived optimal and threshold policies for implementing ART for HNC. Through MC simulation analysis, we demonstrated that the policies obtained from the MDP analysis can significantly outperform fixed-time (one-size-fits-all) policies in terms of both individual toxicity outcomes and the financial burden on the healthcare system. It was discussed that the policies derived from the MDP model allocate resources to patients that need them most, without limiting the number of re-plannings per patient; they can effectively prevent extreme toxicities at an average cost of less than one re-planning per patient. In addition to these results, by providing an evidence-based, resource-aware, and scalable analytical framework for individualized adaptation, we lay the groundwork for future research aimed at developing an online adaptive workflow.

## Data Availability

All data produced are available online at https://github.com/DSuarez03/Cost_Effective_ART_MDP.

## APPENDIX A CHEERS 2022 Checklist

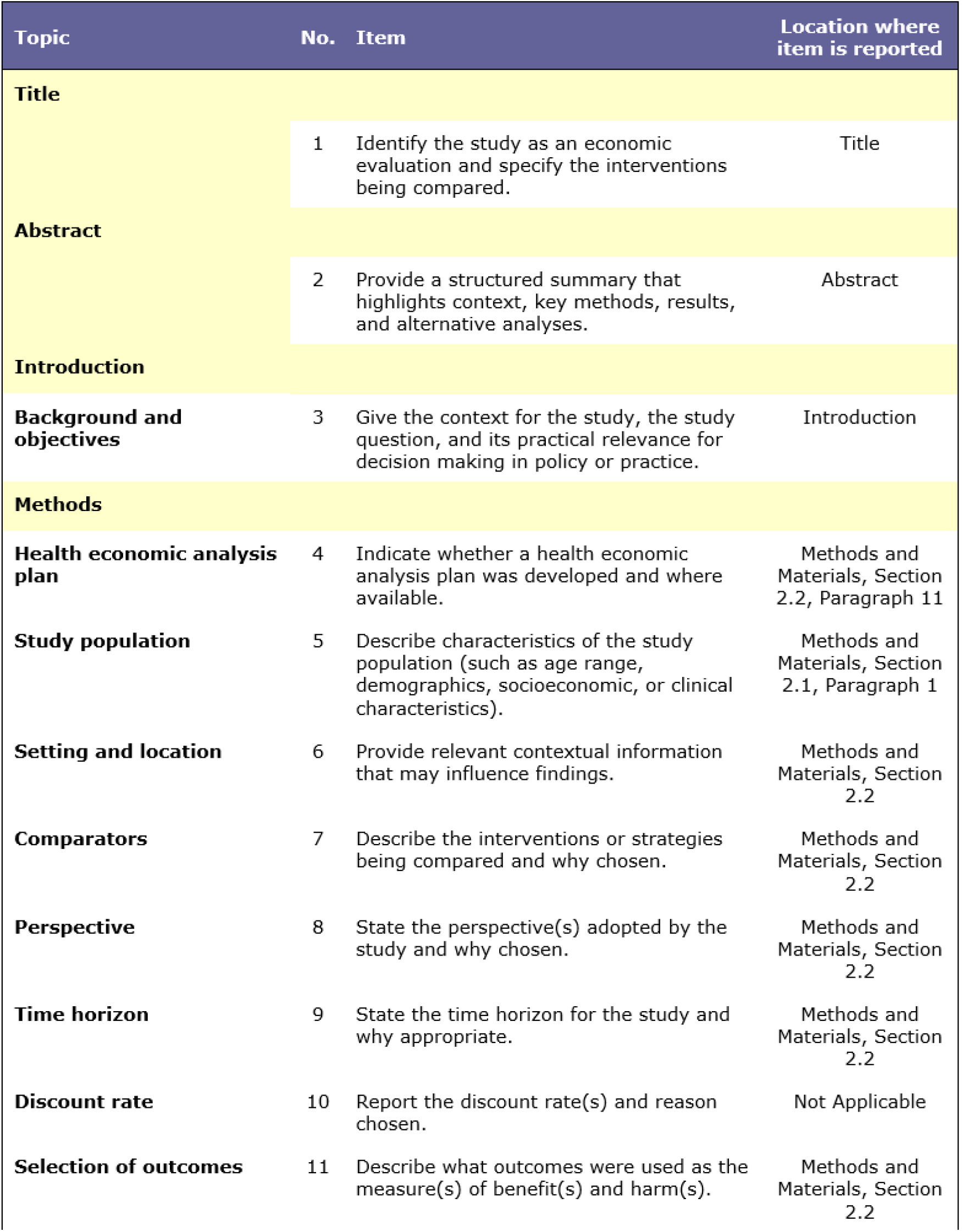
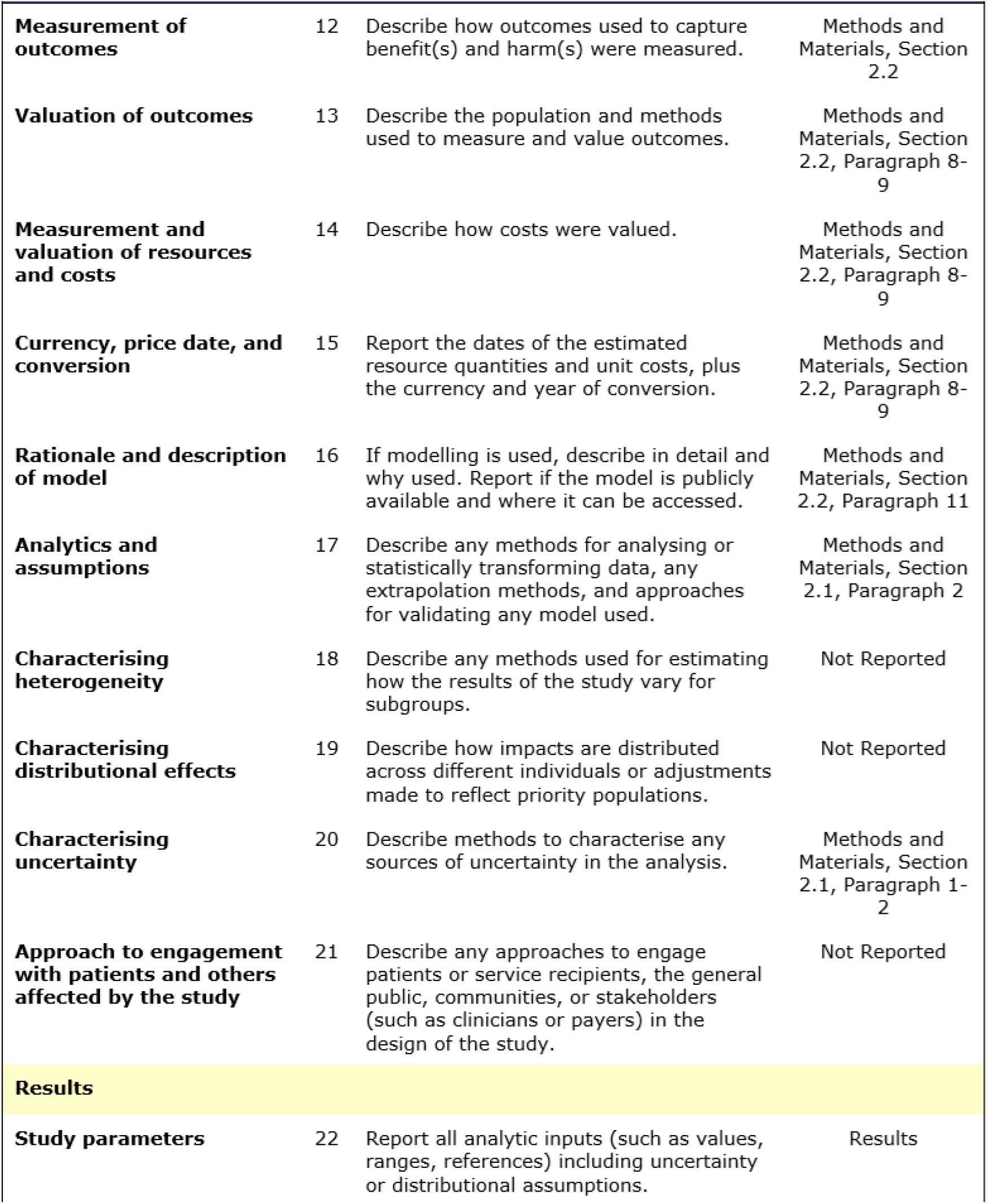
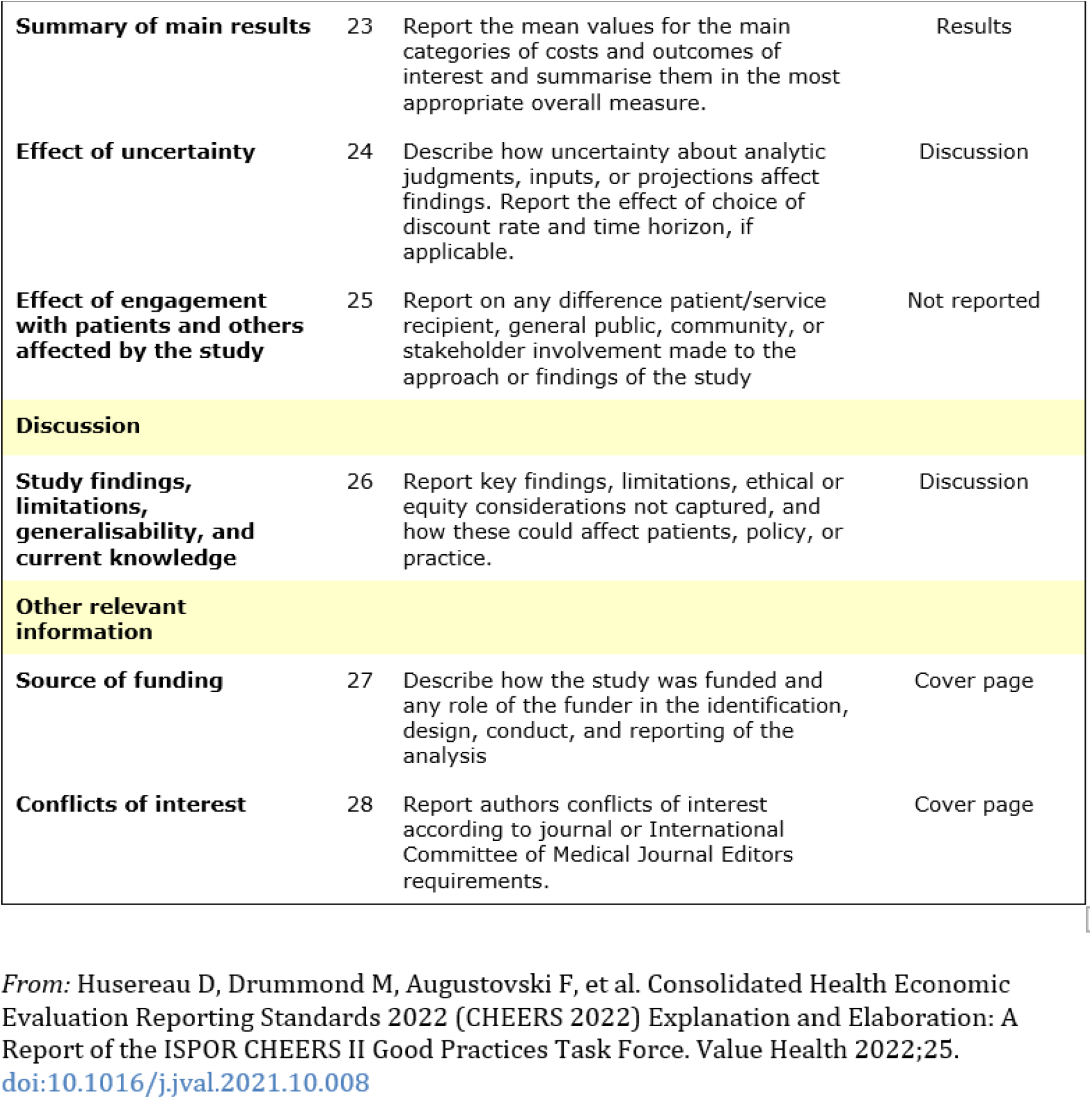

## APPENDIX B Patient Characteristics^1^

| Variable | Number of Patients = 52 | Percentage |
| --- | --- | --- |
| Sex | Male | 69% |
|  | Female | 31% |
| Age | 18-65 years | 88% |
|  | ≥ 65 years | 12% |
| T-classification | Tis-T1 | 8% |
|  | T2 | 15% |
|  | T3 | 13% |
|  | T4 | 54% |
|  | Recurrence | 8% |
|  | Unknown | 2% |
|  | Post-surgery | 23% |
| N-classification | N0 | 27% |
|  | N1 | 19% |
|  | N2 | 38% |
|  | N3 | 2% |
| Primary site | Larynx | 2% |
|  | Oropharynx | 25% |
|  | Oral cavity | 8% |
|  | Hypopharynx | 0% |
|  | Nasopharynx | 29% |
|  | Sinonasal | 23% |
| Pathology | Adenocarcinoma | 2% |
|  | Neuroblastoma | 6% |
|  | Neuroendocrine | 8% |
|  | Squamous cell carcinoma | 50% |
|  | Undifferentiated carcinoma | 17% |
|  | Other | 17% |
| Treatment modality | Radiotherapy alone | 31% |
|  | Induction chemotherapy followed by RT | 4% |
|  | Induction chemotherapy followed by cCRT | 31% |
|  | Concurrent chemoradiation | 27% |
|  | Radiation + Cetuximab | 8% |
| Baseline weight loss | No weight loss | 38% |
|  | Moderate weight loss (1 – 10%) | 33% |
|  | Severe weight loss (>10%) | 6% |
|  | Unknown | 23% |
| Baseline xerostomia | No xerostomia | 4% |
|  | Some xerostomia | 8% |
|  | Unknown | 88% |
| Accelerated RT | Yes | 6% |

Abbreviations: cCRT = concurrent chemoradiation. RT = radiotherapy.
TNM classification according to version 3. Accelerated RT: 2 Gy per fraction, 6 times per week.

## APPENDIX C Transition Probabilities^2^

**Action: No re-planning**

**Table C1:** Transition probabilities from fraction 0 (F0) to fraction 10 (F10) under “no re-planning.”

| $\Delta\text{NTCP @ F10} \rightarrow$<br>$\Delta\text{NTCP @ F0} \downarrow$ | 0% | 1% | 2% | 3% | 4% | 5% | 6% | 7% | 8% | 9% | 10% | 11% | 12% |
| --- | --- | --- | --- | --- | --- | --- | --- | --- | --- | --- | --- | --- | --- |
| 0% | **0.50** | **0.13** | **0.08** | **0.11** | **0.04** | **0.04** | 0.00 | **0.02** | 0.00 | 0.00 | **0.02** | **0.02** | **0.04** |

**Table C2:**
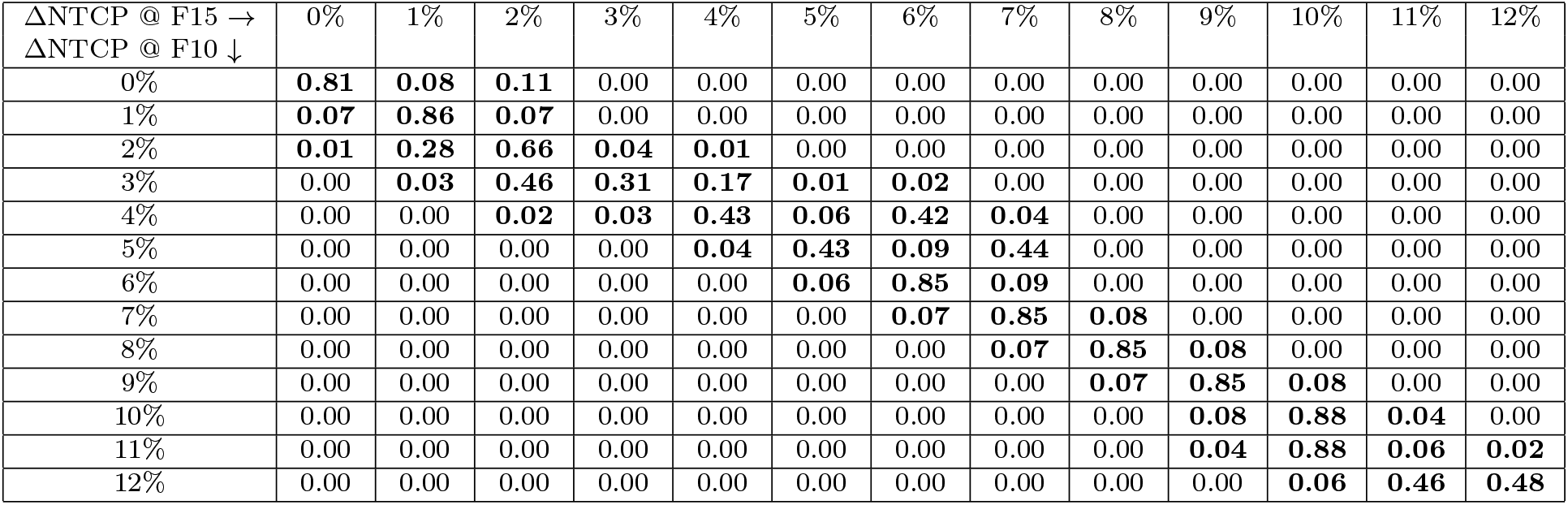
Smoothed transition probabilities from fraction 10 (F10) to fraction 15 (F15) under “no re-planning.” The same probabilities apply to subsequent transitions, i.e., from F15 to F20, from F20 to F25, and from F25 to end-treatment, under “no re-planning.”

**Action: Re-planning**

**Table C3:** Smoothed transition probabilities from fraction 10 to fraction 15 under “re-planning.”

| $\Delta\text{NTCP @ F15} \rightarrow$<br>$\Delta\text{NTCP @ F10} \downarrow$ | 0% | 1% | 2% | 3% | 4% | 5% | 6% | 7% | 8% | 9% | 10% | 11% | 12% |
| --- | --- | --- | --- | --- | --- | --- | --- | --- | --- | --- | --- | --- | --- |
| 0% | **0.81** | **0.08** | **0.11** | 0.00 | 0.00 | 0.00 | 0.00 | 0.00 | 0.00 | 0.00 | 0.00 | 0.00 | 0.00 |
| 1% | **0.81** | **0.08** | **0.11** | 0.00 | 0.00 | 0.00 | 0.00 | 0.00 | 0.00 | 0.00 | 0.00 | 0.00 | 0.00 |
| 2% | **0.07** | **0.86** | **0.07** | 0.00 | 0.00 | 0.00 | 0.00 | 0.00 | 0.00 | 0.00 | 0.00 | 0.00 | 0.00 |
| 3% | **0.07** | **0.86** | **0.07** | 0.00 | 0.00 | 0.00 | 0.00 | 0.00 | 0.00 | 0.00 | 0.00 | 0.00 | 0.00 |
| 4% | **0.07** | **0.86** | **0.07** | 0.00 | 0.00 | 0.00 | 0.00 | 0.00 | 0.00 | 0.00 | 0.00 | 0.00 | 0.00 |
| 5% | **0.01** | **0.28** | **0.66** | **0.04** | **0.01** | 0.00 | 0.00 | 0.00 | 0.00 | 0.00 | 0.00 | 0.00 | 0.00 |
| 6% | **0.01** | **0.28** | **0.66** | **0.04** | **0.01** | 0.00 | 0.00 | 0.00 | 0.00 | 0.00 | 0.00 | 0.00 | 0.00 |
| 7% | **0.01** | **0.28** | **0.66** | **0.04** | **0.01** | 0.00 | 0.00 | 0.00 | 0.00 | 0.00 | 0.00 | 0.00 | 0.00 |
| 8% | 0.00 | **0.03** | **0.46** | **0.31** | **0.17** | **0.01** | **0.02** | 0.00 | 0.00 | 0.00 | 0.00 | 0.00 | 0.00 |
| 9% | 0.00 | **0.03** | **0.46** | **0.31** | **0.17** | **0.01** | **0.02** | 0.00 | 0.00 | 0.00 | 0.00 | 0.00 | 0.00 |
| 10% | 0.00 | **0.03** | **0.46** | **0.31** | **0.17** | **0.01** | **0.02** | 0.00 | 0.00 | 0.00 | 0.00 | 0.00 | 0.00 |
| 11% | 0.00 | 0.00 | **0.02** | **0.03** | **0.43** | **0.06** | **0.42** | **0.04** | 0.00 | 0.00 | 0.00 | 0.00 | 0.00 |
| 12% | 0.00 | 0.00 | **0.02** | **0.03** | **0.43** | **0.06** | **0.42** | **0.04** | 0.00 | 0.00 | 0.00 | 0.00 | 0.00 |

<sup>2</sup>F. Nosrat, C. Dede, L. B. McCullum, R. Garcia, A. S. Mohamed, J. G. Scott, J. E. Bates, B. A. McDonald, K. A. Wahid, M. A. Naser, R. He, A. C. Moreno, L. V. van Dijk, K. K. Brock, J. Heukelom, S. Hosseinian, M. Hemmati, A. J. Schaefer, and C. D. Fuller. Optimal timing of organs-at-risk-sparing adaptive radiation therapy for head-and-neck cancer under re-planning resource constraints. *medRxiv*, 2024. DOI: 10.1101/2024.04.01.24305163.

**Table C4:** Smoothed transition probabilities from fraction 15 to fraction 20 under “re-planning.”

| $\Delta\text{NTCP @ F20} \rightarrow$<br>$\Delta\text{NTCP @ F15} \downarrow$ | 0% | 1% | 2% | 3% | 4% | 5% | 6% | 7% | 8% | 9% | 10% | 11% | 12% |
| --- | --- | --- | --- | --- | --- | --- | --- | --- | --- | --- | --- | --- | --- |
| 0% | **0.81** | **0.08** | **0.11** | 0.00 | 0.00 | 0.00 | 0.00 | 0.00 | 0.00 | 0.00 | 0.00 | 0.00 | 0.00 |
| 1% | **0.07** | **0.86** | **0.07** | 0.00 | 0.00 | 0.00 | 0.00 | 0.00 | 0.00 | 0.00 | 0.00 | 0.00 | 0.00 |
| 2% | **0.07** | **0.86** | **0.07** | 0.00 | 0.00 | 0.00 | 0.00 | 0.00 | 0.00 | 0.00 | 0.00 | 0.00 | 0.00 |
| 3% | **0.01** | **0.28** | **0.66** | **0.04** | **0.01** | 0.00 | 0.00 | 0.00 | 0.00 | 0.00 | 0.00 | 0.00 | 0.00 |
| 4% | **0.01** | **0.28** | **0.66** | **0.04** | **0.01** | 0.00 | 0.00 | 0.00 | 0.00 | 0.00 | 0.00 | 0.00 | 0.00 |
| 5% | 0.00 | **0.03** | **0.46** | **0.31** | **0.17** | **0.01** | **0.02** | 0.00 | 0.00 | 0.00 | 0.00 | 0.00 | 0.00 |
| 6% | 0.00 | **0.03** | **0.46** | **0.31** | **0.17** | **0.01** | **0.02** | 0.00 | 0.00 | 0.00 | 0.00 | 0.00 | 0.00 |
| 7% | 0.00 | 0.00 | **0.02** | **0.03** | **0.43** | **0.06** | **0.42** | **0.04** | 0.00 | 0.00 | 0.00 | 0.00 | 0.00 |
| 8% | 0.00 | 0.00 | **0.02** | **0.03** | **0.43** | **0.06** | **0.42** | **0.04** | 0.00 | 0.00 | 0.00 | 0.00 | 0.00 |
| 9% | 0.00 | 0.00 | 0.00 | 0.00 | **0.04** | **0.43** | **0.09** | **0.44** | 0.00 | 0.00 | 0.00 | 0.00 | 0.00 |
| 10% | 0.00 | 0.00 | 0.00 | 0.00 | **0.04** | **0.43** | **0.09** | **0.44** | 0.00 | 0.00 | 0.00 | 0.00 | 0.00 |
| 11% | 0.00 | 0.00 | 0.00 | 0.00 | 0.00 | **0.06** | **0.85** | **0.09** | 0.00 | 0.00 | 0.00 | 0.00 | 0.00 |
| 12% | 0.00 | 0.00 | 0.00 | 0.00 | 0.00 | **0.06** | **0.85** | **0.09** | 0.00 | 0.00 | 0.00 | 0.00 | 0.00 |

**Table C5:** Smoothed transition probabilities from fraction 20 to fraction 25 under “re-planning.”

| $\Delta\text{NTCP @ F25} \rightarrow$<br>$\Delta\text{NTCP @ F20} \downarrow$ | 0% | 1% | 2% | 3% | 4% | 5% | 6% | 7% | 8% | 9% | 10% | 11% | 12% |
| --- | --- | --- | --- | --- | --- | --- | --- | --- | --- | --- | --- | --- | --- |
| 0% | **0.81** | **0.08** | **0.11** | 0.00 | 0.00 | 0.00 | 0.00 | 0.00 | 0.00 | 0.00 | 0.00 | 0.00 | 0.00 |
| 1% | **0.07** | **0.86** | **0.07** | 0.00 | 0.00 | 0.00 | 0.00 | 0.00 | 0.00 | 0.00 | 0.00 | 0.00 | 0.00 |
| 2% | **0.07** | **0.86** | **0.07** | 0.00 | 0.00 | 0.00 | 0.00 | 0.00 | 0.00 | 0.00 | 0.00 | 0.00 | 0.00 |
| 3% | **0.01** | **0.28** | **0.66** | **0.04** | **0.01** | 0.00 | 0.00 | 0.00 | 0.00 | 0.00 | 0.00 | 0.00 | 0.00 |
| 4% | 0.00 | **0.03** | **0.46** | **0.31** | **0.17** | **0.01** | **0.02** | 0.00 | 0.00 | 0.00 | 0.00 | 0.00 | 0.00 |
| 5% | 0.00 | **0.03** | **0.46** | **0.31** | **0.17** | **0.01** | **0.02** | 0.00 | 0.00 | 0.00 | 0.00 | 0.00 | 0.00 |
| 6% | 0.00 | 0.00 | **0.02** | **0.03** | **0.43** | **0.06** | **0.42** | **0.04** | 0.00 | 0.00 | 0.00 | 0.00 | 0.00 |
| 7% | 0.00 | 0.00 | 0.00 | 0.00 | **0.04** | **0.43** | **0.09** | **0.44** | 0.00 | 0.00 | 0.00 | 0.00 | 0.00 |
| 8% | 0.00 | 0.00 | 0.00 | 0.00 | **0.04** | **0.43** | **0.09** | **0.44** | 0.00 | 0.00 | 0.00 | 0.00 | 0.00 |
| 9% | 0.00 | 0.00 | 0.00 | 0.00 | 0.00 | **0.06** | **0.85** | **0.09** | 0.00 | 0.00 | 0.00 | 0.00 | 0.00 |
| 10% | 0.00 | 0.00 | 0.00 | 0.00 | 0.00 | 0.00 | **0.07** | **0.85** | **0.08** | 0.00 | 0.00 | 0.00 | 0.00 |
| 11% | 0.00 | 0.00 | 0.00 | 0.00 | 0.00 | 0.00 | **0.07** | **0.85** | **0.08** | 0.00 | 0.00 | 0.00 | 0.00 |
| 12% | 0.00 | 0.00 | 0.00 | 0.00 | 0.00 | 0.00 | 0.00 | **0.07** | **0.85** | **0.08** | 0.00 | 0.00 | 0.00 |

**Table C6:** Smoothed transition probabilities from fraction 25 to end-treatment under “re-planning.”

| $\Delta\text{NTCP @ end} \rightarrow$<br>$\Delta\text{NTCP @ F25} \downarrow$ | 0% | 1% | 2% | 3% | 4% | 5% | 6% | 7% | 8% | 9% | 10% | 11% | 12% |
| --- | --- | --- | --- | --- | --- | --- | --- | --- | --- | --- | --- | --- | --- |
| 0% | **0.81** | **0.08** | **0.11** | 0.00 | 0.00 | 0.00 | 0.00 | 0.00 | 0.00 | 0.00 | 0.00 | 0.00 | 0.00 |
| 1% | **0.07** | **0.86** | **0.07** | 0.00 | 0.00 | 0.00 | 0.00 | 0.00 | 0.00 | 0.00 | 0.00 | 0.00 | 0.00 |
| 2% | **0.01** | **0.28** | **0.66** | **0.04** | **0.01** | 0.00 | 0.00 | 0.00 | 0.00 | 0.00 | 0.00 | 0.00 | 0.00 |
| 3% | 0.00 | **0.03** | **0.46** | **0.31** | **0.17** | **0.01** | **0.02** | 0.00 | 0.00 | 0.00 | 0.00 | 0.00 | 0.00 |
| 4% | 0.00 | **0.03** | **0.46** | **0.31** | **0.17** | **0.01** | **0.02** | 0.00 | 0.00 | 0.00 | 0.00 | 0.00 | 0.00 |
| 5% | 0.00 | 0.00 | **0.02** | **0.03** | **0.43** | **0.06** | **0.42** | **0.04** | 0.00 | 0.00 | 0.00 | 0.00 | 0.00 |
| 6% | 0.00 | 0.00 | 0.00 | 0.00 | **0.04** | **0.43** | **0.09** | **0.44** | 0.00 | 0.00 | 0.00 | 0.00 | 0.00 |
| 7% | 0.00 | 0.00 | 0.00 | 0.00 | 0.00 | **0.06** | **0.85** | **0.09** | 0.00 | 0.00 | 0.00 | 0.00 | 0.00 |
| 8% | 0.00 | 0.00 | 0.00 | 0.00 | 0.00 | 0.00 | **0.07** | **0.85** | **0.08** | 0.00 | 0.00 | 0.00 | 0.00 |
| 9% | 0.00 | 0.00 | 0.00 | 0.00 | 0.00 | 0.00 | 0.00 | **0.07** | **0.85** | **0.08** | 0.00 | 0.00 | 0.00 |
| 10% | 0.00 | 0.00 | 0.00 | 0.00 | 0.00 | 0.00 | 0.00 | **0.07** | **0.85** | **0.08** | 0.00 | 0.00 | 0.00 |
| 11% | 0.00 | 0.00 | 0.00 | 0.00 | 0.00 | 0.00 | 0.00 | 0.00 | **0.07** | **0.85** | **0.08** | 0.00 | 0.00 |
| 12% | 0.00 | 0.00 | 0.00 | 0.00 | 0.00 | 0.00 | 0.00 | 0.00 | 0.00 | **0.08** | **0.88** | **0.04** | 0.00 |

## APPENDIX D Terminal Rewards

We consider a patient’s quality of life over *T* years after treatment. Let *A* denote the event that the patient’s lifespan exceeds *T*, and *A*^*c*^ be the complementary event, meaning the patient dies within *T* years post-treatment. We denote the patient’s *T* -year survival probability with *S*, that is *P* (*A*) = *S* and *P* (*A*^*c*^) = 1 *−S*. In the former event, the patient’s quality of life will be *Q <* 1 if they experience a certain radiation toxicity, with the probability *NTCP*. Otherwise, their quality of life will be 1, with the probability 1 *− NTCP*. We naturally assume the quality of life of zero in the latter event. Based on the described chain of conditional probabilities, the patient’s expected quality-adjusted life years (QALY) over *T* years, denoted by 𝔼 QALY, can be calculated as follows:

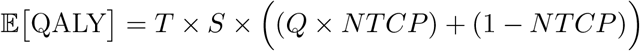

Therefore, the difference in the patient’s QALY resulting from two distinct normal tissue complication probabilities, denoted by *NTCP*_1_ and *NTCP*_2_, can be expressed as follows:

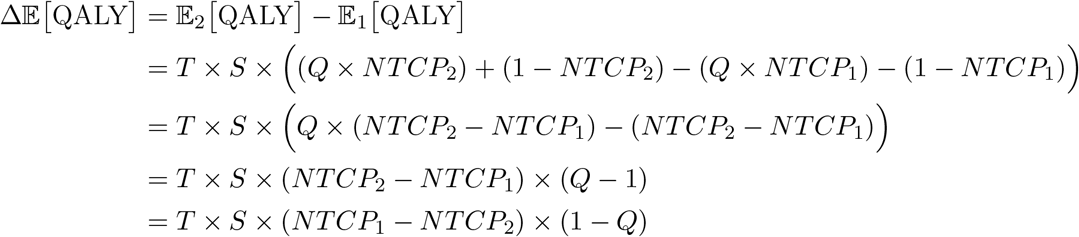

Define Δ*NTCP* = *NTCP*_1_ *− NTCP*_2_, and let *W* denote a willingness-to-pay per QALY ($) value. Thus, the corresponding change in the patient’s expected QALY (over *T* years) can be equivalently expressed in monetary terms as follows:

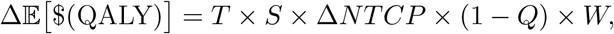

where $(QALY) denotes the monetary equivalence of the patient’s QALY over *T* years. Because *Q <* 1, observe that *NTCP*_2_ *> NTCP*_1_ implies Δ𝔼 $(QALY)] < 0.

## APPENDIX E Quality-of-Life Factors^3^

| Endpoint | Quality-adjustment factor | Reference |
| --- | --- | --- |
| Xerostomia<br>(grade $\geq 2$) | 0.827 (range: 0.819 – 0.841)<br>0.817 ± 0.19 | Kohler et al. 2013 (PMID: 24138916)<sup>4</sup><br>Ramaekers et al. 2011 (PMID: 21683647)<sup>5</sup> |
| Dysphagia<br>(grade $\geq 2$) | 0.85 ± 0.11<br>0.803 ± 0.14 | Rudmik et al. 2015 (PMID: 26171771)<sup>6</sup><br>Ramaekers et al. 2011 (PMID: 21683647)<sup>5</sup> |

<sup>4</sup>R. E. Kohler, N. C. Sheets, S. B. Wheeler, C. Nutting, E. Hall, and B. S. Chera. Two-year and lifetime cost-effectiveness of intensity modulated radiation therapy versus 3-dimensional conformal radiation therapy for head-and-neck cancer. *International Journal of Radiation Oncology, Biology, Physics*, 87(4):683–689, 2013.
<sup>5</sup>B. L. Ramaekers, M. A. Joore, J. P. Grutters, P. Van Den Ende, J. De Jong, R. Houben, P. Lambin, M. Christianen, I. Beetz, M. Pijls-Johannesma, et al. The impact of late treatment-toxicity on generic health-related quality of life in head and neck cancer patients after radiotherapy. *Oral oncology*, 47(8):768–774, 2011.
<sup>6</sup>L. Rudmik, W. An, D. Livingstone, W. Matthews, H. Seikaly, R. Scrimger, and D. Marshall. Making a case for high-volume robotic surgery centers: A cost-effectiveness analysis of transoral robotic surgery. *Journal of Surgical Oncology*, 112(2):155–163, 2015.

## Footnotes

1 J. Heukelom, M. E. Kantor, A. S. Mohamed, H. Elhalawani, E. Kocak-Uzel, T. Lin, J. Yang, M. Aristophanous, C. R. Rasch, C. D. Fuller, and J. Sonke. Differences between planned and delivered dose for head and neck cancer, and their consequences for normal tissue complication probability and treatment adaptation. Radiotherapy and Oncology, 142:100–106, 2020.

2 F. Nosrat, C. Dede, L. B. McCullum, R. Garcia, A. S. Mohamed, J. G. Scott, J. E. Bates, B. A. McDonald, K. A. Wahid, M. A. Naser, R. He, A. C. Moreno, L. V. van Dijk, K. K. Brock, J. Heukelom, S. Hosseinian, M. Hemmati, A. J. Schaefer, and C. D. Fuller. Optimal timing of organs-at-risk-sparing adaptive radiation therapy for head-and-neck cancer under re-planning resource constraints. *medRxiv*, 2024. DOI: 10.1101/2024.04.01.24305163.

3 N. P. Brodin, R. Kabarriti, M. Pankuch, C. B. Schechter, V. Gondi, S. Kalnicki, C. Guha, M. K. Garg, and W. A. Tomé. A quantitative clinical decision–support strategy identifying which patients with oropharyngeal head and neck cancer may benefit the most from proton radiation therapy. *International Journal of Radiation Oncology, Biology, Physics*, 104(3):540–552, 2019.

4 R. E. Kohler, N. C. Sheets, S. B. Wheeler, C. Nutting, E. Hall, and B. S. Chera. Two-year and lifetime cost-effectiveness of intensity modulated radiation therapy versus 3-dimensional conformal radiation therapy for head-and-neck cancer. *International Journal of Radiation Oncology, Biology, Physics*, 87(4):683–689, 2013.

5 B. L. Ramaekers, M. A. Joore, J. P. Grutters, P. Van Den Ende, J. De Jong, R. Houben, P. Lambin, M. Christianen, I. Beetz, M. Pijls-Johannesma, et al. The impact of late treatment-toxicity on generic health-related quality of life in head and neck cancer patients after radiotherapy. *Oral oncology*, 47(8):768–774, 2011.

6 L. Rudmik, W. An, D. Livingstone, W. Matthews, H. Seikaly, R. Scrimger, and D. Marshall. Making a case for high-volume robotic surgery centers: A cost-effectiveness analysis of transoral robotic surgery. *Journal of Surgical Oncology*, 112(2):155–163, 2015.

